# Assortative mating across nine psychiatric disorders is consistent and persistent over cultures and generations

**DOI:** 10.1101/2024.09.19.24314024

**Authors:** Chun Chieh Fan, Saeid Rasekhi Dehkordi, Richard Border, Lucy Shao, Bohan Xu, Robert Loughnan, Wesley K. Thompson, Le-Yin Hsu, Mei-Chen Lin, Chi-Fung Cheng, Rou-Yi Lai, Mei-Hsin Su, Wei-Yi Kao, Thomas Werge, Chi-Shin Wu, Andrew J. Schork, Noah Zaitlen, Alfonso Buil Demur, Shi-Heng Wang

## Abstract

Emerging evidence has shown that assortative mating (AM) is a key factor that shapes the landscape of complex human traits. It can increase the overall prevalence of disorders, influence occurrences of comorbidities, and bias estimation of genetic architectures. However, there is lack of large-scale studies to examine the cultural differences and the generational trends of AM for psychiatric disorders. Here, using national registry datasets, we conduct the largest scale of AM analyses on nine psychiatric disorders, with up to 1.4 million mated cases and 6 million matched controls. We performed meta-analyses on AM estimates from Taiwan, Denmark, and Sweden, to examine the potential impact of cultural differences. Generational changes for people born after 1930s were investigated as well. We found that AM of psychiatric disorders are consistent across nations and persistent over generations, with a small proportion of disorders showing generational changes of AM. Our results provide additional insight into the mechanisms of AM across psychiatric disorders and have evident implications on the estimation of the genetic architectures of psychiatric disorders.

## Introduction

Assortative mating (AM), a non-random pairing process in which individuals with similar phenotypes mate more frequently than would be expected, has long been considered as ubiquitous across traits and universal in human society ^1,2^. It can be driven by the shared environment between spouses that makes them more alike, or the social homogamy that constrains the choices of spouses based on traits. It can also be the end results of the active assortment that people are attracted to the mate with similar traits. While some of those driving forces are more sensitive to the cultural changes, the cumulative effect of the AM over generations can lead to evident impact on the trait variations and disease prevalences ^1–11^. Therefore, investigating the patterns of AM is important for understanding and modeling the population dynamics of human complex traits.

This is especially critical for psychiatric disorders. The tendency to share the same diagnoses between spouses have long been reported ^1,3–5^. However, the number of large-scale studies on the AM of psychiatric disorders are limited. Based on the Swedish registry, Nordsletten et al. reported substantial amount of AM for pairs with the same psychiatric disorders in either Attention Deficits Hyperactivity Disorder (ADHD), Autism Spectrum Disorder (ASD), Schizophrenia (SCZ), and Substance Use Disorder (SUD) ^12^. Though including fewer psychiatric disorders, a study examining the cross-trait AM also found evident spousal correlations across psychiatric disorders in Denmark ^13^. It is unclear if these patterns hold beyond Nordic countries. A recent analyses on physical traits based on Japan Biobank and UKBiobank found regional differences of AM ^14^. Physical activities, food preferences, diabetes, and coronary artery diseases have evident regional variations, indicating the potential involvement of social homogamy for cultural sensitive practices ^14^. Given the complexity of psychiatric disorders, it is possible behaviors would differ across cultures, leading to differences in observed AM. For example, in an environment with less social stigma of the psychiatric disorders, the mating choices of the patients can be more diverse, reducing the observed AM of the psychiatric disorders accordingly.

Furthermore, there is no investigation on the generational change of the AM for psychiatric disorders. An exogeneous cause to the AM, such as social stigma induced limitations on spousal choices, would not be hold constant over time due to the evolution of psychiatric cares in the past decades. Some argued that the AM of psychiatric disorders is unlikely to persist over generations due to rapid changes in modern society ^11^, hence unlikely to have substantial impact on the heritability and prevalence of the psychiatric disorders. By examining the generational change of observed AM, we can gain insight the contributing factors of spousal resemblance among patients with psychiatric disorders, such as social homogamy, and obtain a crucial piece of information on the degree and dynamics of AM to better assess its impact on current descriptions of the genetic architecture of psychiatric disorders.

To address the aforementioned gap in our knowledge, here, we present the largest scale investigation on AM of psychiatric disorders. The total number of mated cases across cohorts range from 1.4 million (Major Depressive Disorder) to 31,195 (Anorexia Nervosa). The number of matched controls reached up to 6 million spousal pairs. The unprecedented inclusion of 14.8 million mated individuals, enriched for psychiatric disorders, allow us to investigate the relevancy of cultural differences and generational changes. We estimate the cross-trait AM among 9 psychiatric disorders using Taiwan National Health Insurance Research Database (NHIRD), which include individuals born between 1930s to 1990s, and then perform meta-analyses with the estimates derived from two large-scale Nordic registries. We tested the heterogeneity in the estimated spousal cross-traits resemblance to examine the cultural (Nordic versus Taiwan) and generational (birth cohorts) differences in the AM of psychiatric disorders.

## Methods

### Taiwan NHIRD

The NHIRD was derived from Taiwan’s single-payer National Health Insurance program, which covers 99% of the 23 million Taiwan population. Each individual has a unique number as personal identifier, allowing linkage to the records of medical utilization and the diagnostic registrations. Details on how the familial relationships were determined and how the disease status were ascertained can be found in Wang et al, 2022 ^15^. In short, we utilized the dependent policy of Taiwan National Insurance to infer familial relationships, as only spouses and first-degree relatives are eligible as dependents of an insured person. All registry entries from 1997 to 2018 are included to identify unique potential participants and then familial relationships were established based on dependencies across all entries. The relationship records were validated through the maternal/child health registries within NHIRD. The accuracy for the inferred familial relationship is 99.35% ^15^. The legal marriage age of Taiwan without the need for parental consent is 18 years old, hence we include only those individuals who were born between 1930 and 1999 to allow for at least 18 years of the lead time. We exclude same-sex marriage couples, as their legal status was gradually established between 2017 and 2019 in Taiwan. Individuals who have multiple spouses registered were also excluded. The final number of eligible pairs was 4,954,870.

The disorder status is defined as those who ever have diagnostic entries of Schizophrenia (SCZ), Attention Deficit – Hyperactivity Disorder (ADHD), Autism Spectrum Disorder (ASD), Major Depressive Disorder (MDD), Bipolar Disorder (BPD), Anxiety Disorders (Anxiety), Obsessive Compulsive Disorder (OCD), Substance Use Disorder (SUD), or Anorexia Nervosa (AN), for at least one inpatient or two outpatient visits. ICD codes were used to ascertain the diagnostic entries, which are detailed in **Supplementary Table 1**. After intersecting with the familial relationships, we ascertained 79,551 probands with SCZ, 218,867 probands with BPD, 1,005,055 probands with MDD, 1,091 probands with ASD, 10,436 probands with ADHD, 592,245 probands with SUD, 32,914 probands with OCD, 654,661 probands with Anxiety, and 1,615 probands with AN.

After the diagnostic status were ascertained, we utilized the matched case-control design to construe the final analytic samples. This is the same strategy used in the large-scale study of AM in Sweden ^12^. We start with the probands who have the diagnosis of interest, and then randomly selected 5 population controls matched by sex, birth year, calendar year, and living areas. The proportions of all nine psychiatric disorders among the spouses of the ascertained probands and their matched controls were derived, forming the basis for calculating spousal correlations. We choose tetrachoric correlation as the metric for AM because it is known to be insensitive to the prevalence differences, providing a common unit for comparing cultural and generational trends ^16^. To examine the generational effects, we stratify the ascertained probands and matched controls into six different birth-year groups, each 10 years apart starting from 1930. The same procedures were applied to each strata to obtained spousal correlations of each birth-year strata. The sample size of each disorder pair, as well as the proportion of disorders among spouses, can be found in in **Supplementary Table 2 and Suplementary Table 3.**

### Danish National Registry

The Danish Civil Register system has recorded the country’s legal residents since 1968. Each individual is identified through a Civil Register number (CPR number) and is linked to his legal parents, which allows building the Danish genealogy ^17^. The CPR, serves as a crucial connection to the health register that contains hospital visits from 1969. By April 2017, the system had recorded data on over 9 million individuals. The inclusion of psychiatric disorders in the registration system began in 1969. Psychiatric disorder records initially limited to inpatient cases, expanded in 1995 to include outpatient diagnoses ^18^. We extracted individuals born after 1969, encompassing 571,534 couples. The term “couples” refers specifically to those officially registered as such, either through marriage or partnership, with partners of the same sex excluded from this study. Psychiatric disorders were categorized according to the ICD-8 coding system initially, transitioning to ICD-10 in 1995. We eventually ascertained 5,976 cases with SCZ, 5,999 cases with BPD, 54,728 cases with MDD, 1,487 cases with ASD, 10,964 cases with ADHD, 77,187 cases with SUD, 5,809 cases with OCD, 5,166 cases with Anxiety, and 10,103 cases with AN. See **Supplementary Table 4** for further details. Assessing assortative mating in psychiatric disorders has been calculated using tetrachoric correlation within a case-control database, with each case matched to five controls of similar gender and age profiles.

### Sweden Assortative Mating Estimates

The AM estimations from the Swedish cohort are obtained from the published work by Nordsletten et al ^12^. The study was a matched case-control design with one-to-five proband-to-control ratio, matching on age, sex, and county of residence. In total, 707,263 probands were ascertained given the eligible psychiatric diagnoses. Detailed numbers of the case and matched controls can be found in the **Supplementary Table 5**.

### Heritability and co-heritability estimates

For comparison purpose, we estimates the heritability and co-heritability across all nine psychiatric disorders using LD score regression (LDSC) ^19^. The source for the summary statistics of the genome-wide association studies (GWAS) for each of the nine psychiatric disorders are listed in the **Supplemental Table 6**, including SCZ ^20^, ADHD ^21^, ASD^22^, MDD^23^, BPD^24^, Anxiety^25^, OCD^26^, SUD^27^, or AN ^28^.

### Statistical Analysis

Tetrachoric correlations were calculated using the *psych* package in R. We used the *metafor* package to perform random effects meta-analysis and the meta-regressions ^29^. Mean estimates, moderating effects, and the corresponding confidence intervals are obtained via REML. The models are selected based on Akaike Information Criterion. The significance is set with Bonferroni correction, either at experiment-wide level (9*9 possible pairs of disorders) or at study-wide level (81 possible pairs times 4 different comparisons, i.e. Male versus Female, Taiwan versus Nordic). The results are visualized using *ggplot* and *corrplot*.

## Results

### AM across psychiatric disorders in Taiwan

For nine included psychiatric disorders and their 81 possible pairs, the estimations based on female probands and male probands were largely consistent, except those having very rare incidences (Figure 1a-b). All spousal correlations, regardless of the disease pairs, are positive. The concordance to the estimates from the Nordic countries (Denmark and Sweden) are high, with a few higher than the predictive range based on meta-regressions. Estimates based on Taiwan male probands have higher than expected correlations in AN, OCD, and BPD (Figure 1a). Estimates based on Taiwan female probands have higher than expected correlations in OCD and BPD (Figure 1b). Using Taiwan versus Nordic as the fixed moderating effect in a joint mixed effects meta-analysis, we formally tested the differences between AM estimates. Like the meta-regression results, AN, OCD, and BPD have significantly higher spousal correlations among Taiwanese than those derived from Nordic countries (Figure 1c). Meanwhile, other psychiatric disorder pairs have limited evidence for cultural differences. Even for those pairs that have significant differences in AM across countries, the mean spousal correlations across disorders are all significantly greater than zero (Supplementary Figure 1).

**Figure 1.**
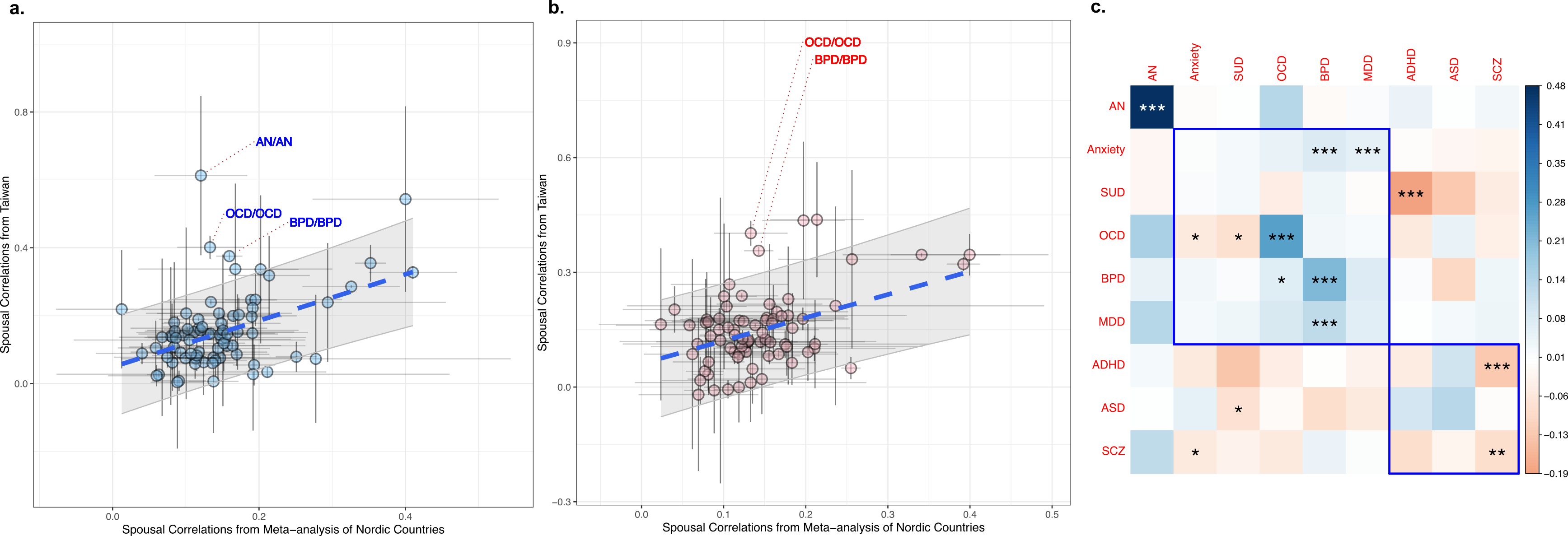
Spousal correlations across nine psychiatric disorders, estimates from Taiwan versus those from Nordic countries (Denmark and Sweden). (a). Estimates based on male proband, comparing to the mean estimates from the Nordic countries. The 95% confidence intervals are plotted for each point estimate and the group regression trend. (b). Estimates based on female proband, comparing to the mean estimates from the Nordic countries. (c). Estimated differences in spousal correlations between Nordic countries and Taiwan. The disease labels are ordered according to the first principal components of the pairwise spousal correlations, while the clusters were highlighted in blue. (* FDR significant with q=0.05; ** Bonferroni correction significant, p < 0.05/81; *** Study-wide significance, p < 0.05/324)

### Generational changes of AM in Taiwan

Because the large sample size and high coverage of records in Taiwan, we can further stratify the cohort according to their birth years while maintaining stable estimates of the spousal correlations. Because the tetrachoric correlation is insensitive to the differences in the prevalences across birth cohorts, as further supported by our simulations (Supplementary Materials; Supplementary Figure 2), the trends of spousal correlations can reveal the generational changes of the AM. Starting from the 1930s until 1990s, the samples were separated into birth cohorts for every 10 years. We then test the significance of the 10-yearly trends, the results are shown in the Figure 2. Figure 2a showcase the point estimates of the average change per 10 years, based on female probands, and their corresponding p values from the mixed effects model. The estimates based on the male probands are shown in the Figure 2b. Of 81 disorder pairs, 16 show study-wide significant trends where all, except for OCD-OCD, has increasing spousal correlations over the years analyzed. Multiple pairs involving SUD shows significant trends upward, with slight differences between females and males (Figure 2a-c). In contrast, the OCD-OCD evidently trends downward for both female and male probands (Figure 2d). Sex specific effects on the generational changes are limited and may reflect the complex generational patterns that are not fully captured by the linear models (Figure 2e). It is worth to note that the AM of ADHD has the largest downward trend over the decades, though the test for trends is not significant, partly due to lack of good estimates from 1930s to 1950s (Figure 2a, b, and f). Those trends cannot be explained by the base rate differences, as the tetrachoric correlation is insensitive to the base rate and the prevalence per strata do not follow the general trends shown here (**Supplementary Materials, Supplementary Figure 2**). The estimates, though, might still be sensitive to the differential censoring over the generations, i.e. those who have the diagnoses were missed in different rate over the generations (**Supplementary Materials, Supplementary Figure 3**). However, differential censoring would only reduce the magnitude of estimated AM via the tetrachoric correlations. The correlations trend upward over generations imply the actual AM can be higher, if the censoring is in place. Meanwhile, censoring cannot explain the correlations trending downward as it is unlikely the ascertainment of OCD and ADHD were getting worse in recent decades.

**Figure 2.**
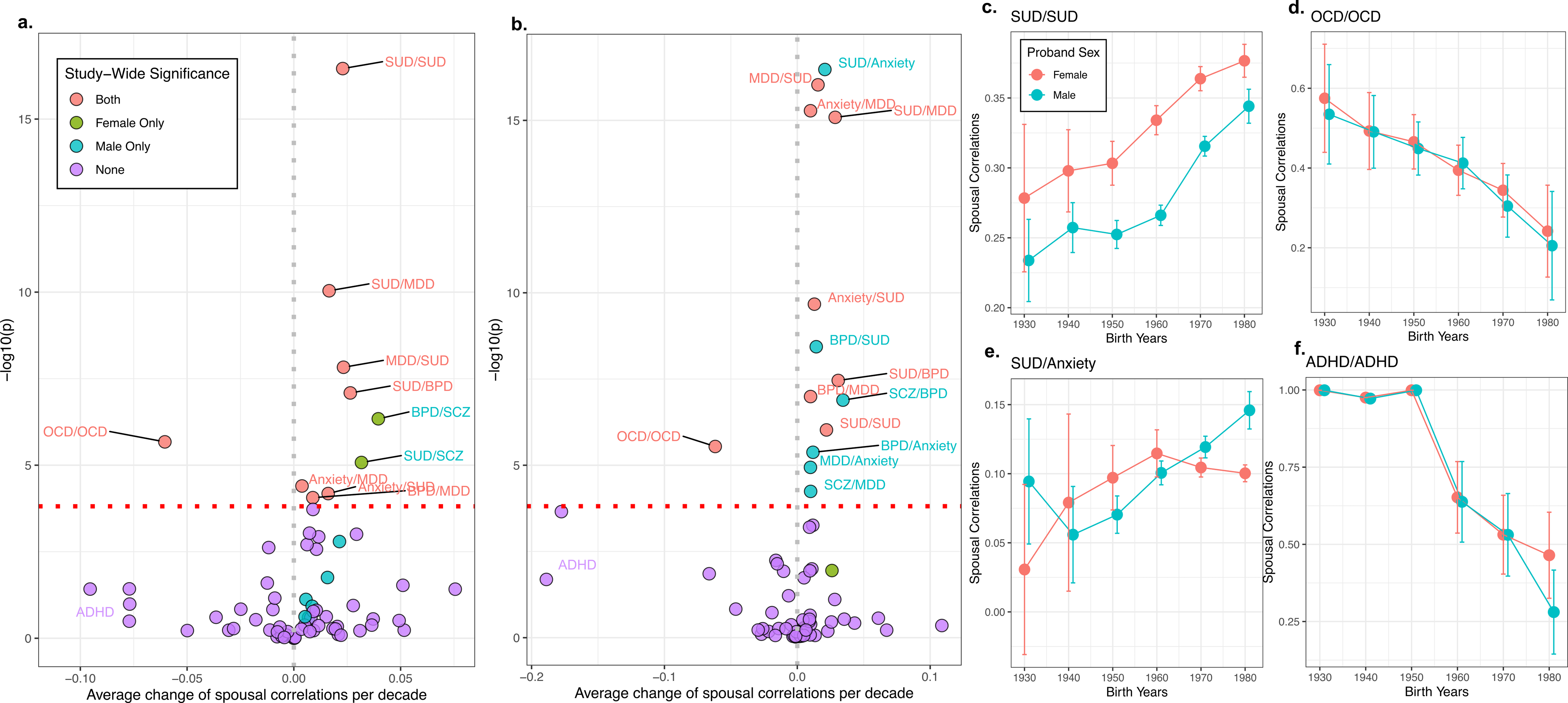
Generational trends of spousal correlations for psychiatric disorders. (a) Volcano plot of the test for generational trends given estimates from female probands over seven birth cohorts since 1930s. The x-axis specifies the point estimates of the average change per 10 years while the y-axis represent the -log10 of the corresponding p-values. Those surpass the study-wide significant threshold are labeled, while the coloring indicates which context they become significance or not. (b) Volcano plot of the test for generational trends given estimates from male probands over seven birth cohorts since 1930s. (c – f) Examples of the generational trends of spousal correlations.

### Impact of the persistent AM across generations

Theoretically, AM would increase the similarities between parent and offspring. Here, we used the unique opportunity provided by the birth cohorts of Taiwan to investigate the correspondence between AM and parent-offspring resemblance over the generations. For each birth cohort, we retained those who have offsprings in the NHIRD and ascertained their diagnoses accordingly. We then calculated the tetrachoric correlations between parents and offsprings, given their diagnostic status, which are shown in the Figure 3. The generational trends of the parent-offspring correlations are similar to the generational trends of the AM. ADHD and OCD are trending downwards whereas all except AN are trending upwards. We also examined the risk differences for the offsprings between those who only have one parent with the disorders and those who have two parents with the disorders. We observed the same increasing trends for MDD, SCZ, and SUD (**Supplementary Figure 4**).

**Figure 3.**
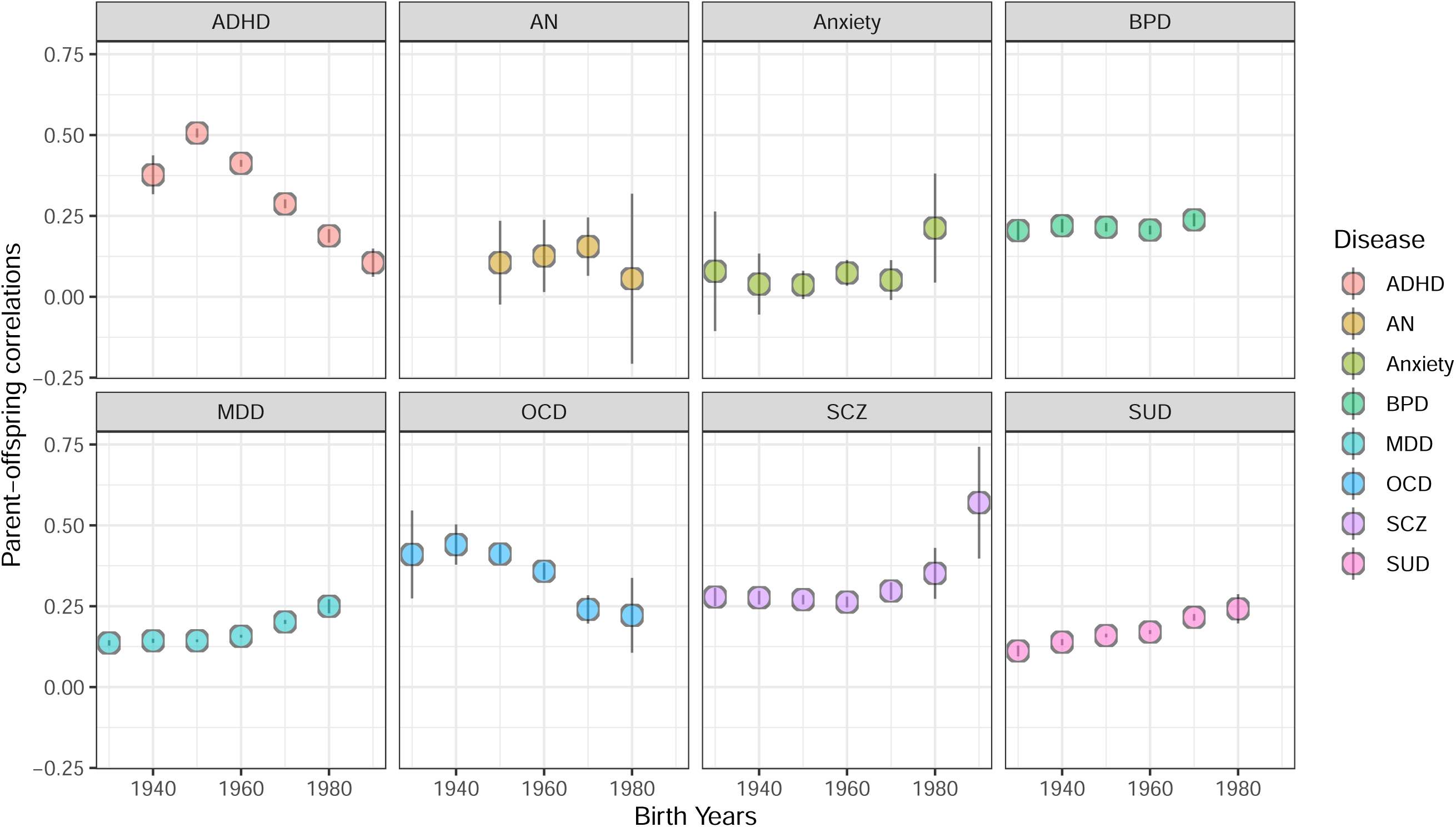
Generational trends of parent-offspring correlations. The estimations were based on the tetrachoric correlations among parent-offspring pairs from the entire cohort given the disease status specified in the figure. The error bars are 95% confidence intervals of the given estimates.

Since the parent-offspring correlations theoretically contain both genetic and rearing effects, we then further examine the genetic covariance by comparing the estimated AM from Taiwanese cohort with the heritability and genetic correlation estimated from genome-wide association studies (GWAS) ^20–28^.

Despite the GWAS were conduct on the population with European ancestry, the observed spousal correlations from the Taiwan samples are highly correlated with the genetic architectures of the examined pairs (Figure 4 a and b). Genetic covariance based on GWAS can explain 48.75% and 56.58% of the observed spousal correlations ascertained from female proband and male proband, respectively. This implicates a shared consequence of a culturally and generationally persistence AM on the population dynamics of psychiatric disorders.

**Figure 4.**
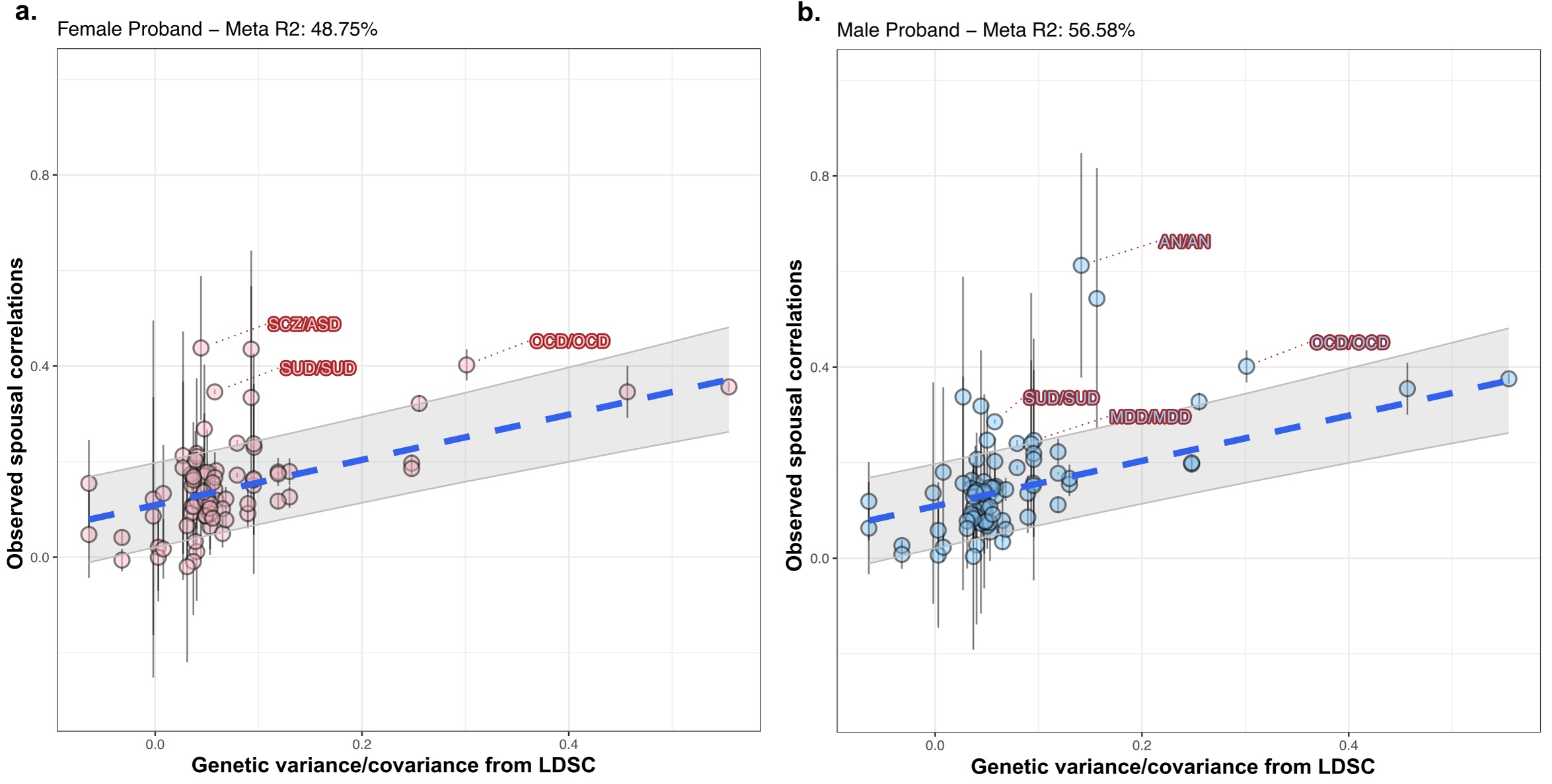
Distribution of the spousal correlations as a function of GWAS based genetic covariance. The blue dotted lines are the linear predictors based on the mixed effects meta-regressions, using estimated genetic variance/covariance as the fixed effect. Gray bands are the 95% confidence intervals of the predicted values. Error bars are the 95% confidence intervals of the AM estimates based on Taiwanese cohort.

## Discussion

Here we have conducted the largest study on AM patterns across psychiatric disorders, with up to 1.4 million cases in probands and 6 million matched controls. Across 9 psychiatric disorders, there is evident positive correlations among spouses, independent of country and generation for the past 90 years. Despite differences in cultural realms, the patterns of AM across psychiatric disorders have limited variations between Taiwan and Nordic countries. Only handful of psychiatric disorders show apparent trends according to birth years, indicating the persistence of AM across time.

Our results have significant implications on our understanding of the genetic architecture of psychiatric disorders. As our results shown, spousal resemblance within and between psychiatric disorder pairs are consistent across countries and persistent throughout the generations, indicating an universal phenomenon. This can have substantial impact on the inference solely based on the estimated genetic correlations, as the persistent AM over generations would conflate the genetic variance with rearing effects while rendering none-causally related traits genetically correlated in the population level ^11,13,30^. Congruent with this notion, we found that the parent-offspring correlations have the same trends as the generational changes of AM. The co-occurrences of the disorders between parents and their offsprings increase over time as the AM persisted for Anxiety, BPD, MDD, OCD, SCZ, and SUD. When the AM evidently decreased over time, such as OCD and ADHD, the parent-offspring correlations also dropped. Meanwhile, we observed high correlations between observed spousal correlations and estimated genetic variance/covariance based on GWAS from individuals with European ancestry, despite completely differences in the study demographics. Those provide converging evidence indicating the cumulative effects of the AM in psychiatric disorders. Yet, it also means that the systematic inflations on genetic effects would be hard to mitigate by simply comparing results from different countries, as AM difference appear to be minimal in our cross-culture analysis. Establishing full pedigree and mating relationships might help to tease apart the conflated effects between AM and genetic inheritance ^31^.

Several disorder pairs involving SUD show upward trends over generations. While estimates without stratification from Taiwan are highly consistent with the estimates from Nordic countries, we cannot determine if Nordic countries also hold such upward trends over time due to lack of available data. Nevertheless, they might reflect a more societal sensitive process given substantial socioeconomical changes in the past half century in Taiwan. While the modern psychiatric service and psychiatric epidemiology in Taiwan has long historical root tracing back to colonial era, the National Health Insurance for universal coverage across Taiwan happened in 1995, greatly reduce the financial burden of the medical cares. The first democratic presidential election happened in 1996. Attitudes towards substance use have been changing over time, leading to active assortment due to the social use of the substances or more willingness to report the substance use problems ^32^.

AM observed for OCD has a unique pattern when compared to other disorder pairs. The estimations from Taiwan are higher than expected given the heritability from GWAS and are significantly higher than those observed in Nordic countries (Figure 1c). Meanwhile, it exhibits apparent generational trends downward (Figure 2). This suggests the force driving the spousal correlations specific to OCD is weakening over generations, bringing it to a level similar to those observed in Nordic countries. The generational change may also explain some variable results of AM estimations of OCD from Swedish registry ^16^. It remains to be seen how these differences will impact the inheritance patterns and the overall prevalence of OCD in the population.

There are limitations in our study. First, the psychiatric probands in our study are all prevalent cases where the diagnoses can happen after the mating. Therefore, shared environment or clinicians’ diagnostic preference toward the same family members might contribute to the observed correlations instead of active assortment. Nonetheless, the consistency of estimates across methods, countries, and generations makes our results unlikely to be solely driven by the differences in the diagnostic practice or social environment. Second, we did not investigate the generational changes of AM in the Nordic countries due to data availability. The converging results on the marginal estimates across Taiwan, Denmark, and Sweden do not guarantee the same observation on the generational trends will hold true. It would be of interest to further explore the birth cohorts in the countries other than Taiwan.

Given aforementioned caveats, our results indicate the patterns of AM across psychiatric disorders are consistent and persistent. This observed phenomenon can contribute to the co-occurrence of the psychiatric disorders and bias genetic estimates of two biologically unrelated but assorted disorders. Given the ubiquitousness of the AM, it is important to take non-random mating patterns into consideration when designing genetic studies of psychiatric disorders.

## Data Availability

All data produced in the present work are contained in the manuscript. For the raw data, the NHIRD used in this study is held by the Taiwan Ministry of Health and Welfare and under controlled access. Researchers interested in accessing the data set can apply form to the Ministry of Health and Welfare requesting access.

## Acknowledgement

This work was supported by National Health Research Institutes (NHRI-EX109-10931PI, NHRI-EX110-10931PI, NHRI-EX111-10931PI; SHW). CCF is supported by NIH R01 MH122688 and R01 MH128959.

## Data Availability

The NHIRD used in this study is held by the Taiwan Ministry of Health and Welfare and under controlled access. Researchers interested in accessing the data set can apply form to the Ministry of Health and Welfare requesting access.

