## Supplementary Materials for "Assortative mating across nine psychiatric disorders is consistent and persistent over cultures and generations"

**Title:**

Supplementary Materials
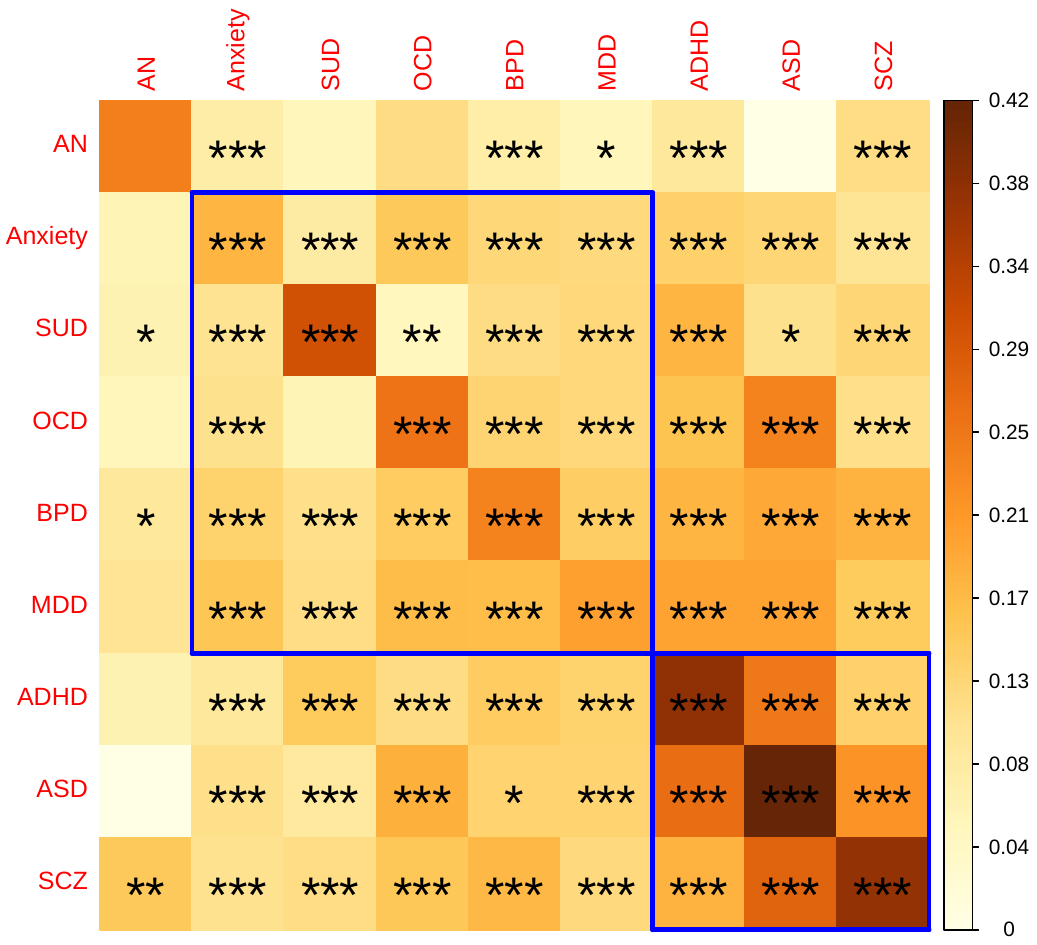

Supplementary Figure 1. Mean spousal correlations across nine psychiatric disorders, based on the mixed effects meta-analyses of estimates from Taiwan, Denmark, and Sweden. (* FDR significant with q=0.05; ** Bonferroni correction signifiant, p < 0.05/81; *** Study-wide signifiance, p < 0.05/324). The disease labels are ordered according to the first principal components of the pairwise spousal correlations, while the clusters were highlighted in blue.

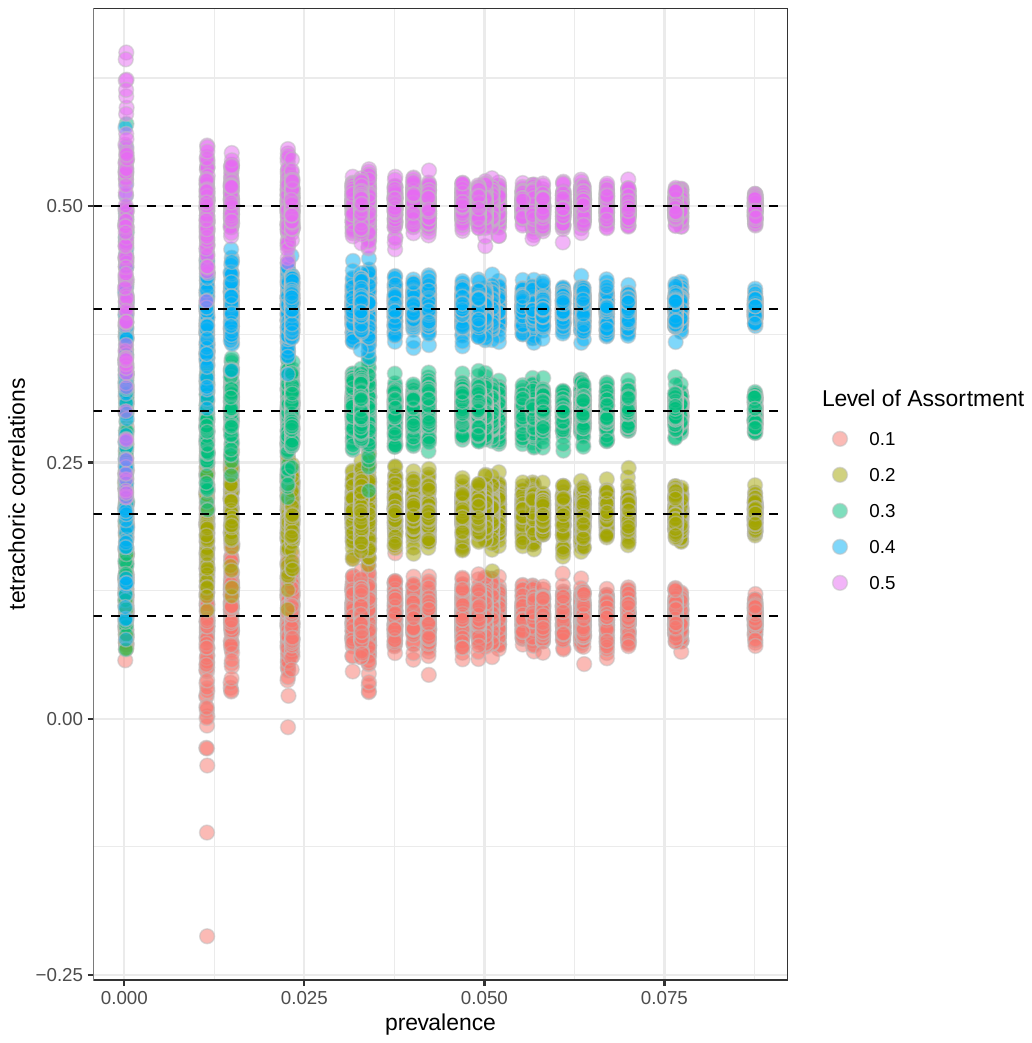

Supplementary Figure 2. Simulations on the impact of prevalence on the estimations of AM with tetrachoric correlation. The total cohort size is fixed at 10,000 pairs while the age of the individuals are assigned as uniformly random. The prevalence per age strata is assigned accordingly. We varied five different levels of assortment and repeated the simulations 100 times per parameter scenario. The result shows that the point estimate of the tetrachoric correlations is unbiased and invariant to the base rate (prevalence). Lower prevalence would only lead to higher sampling variance because reduced number of cases in a closed cohort.

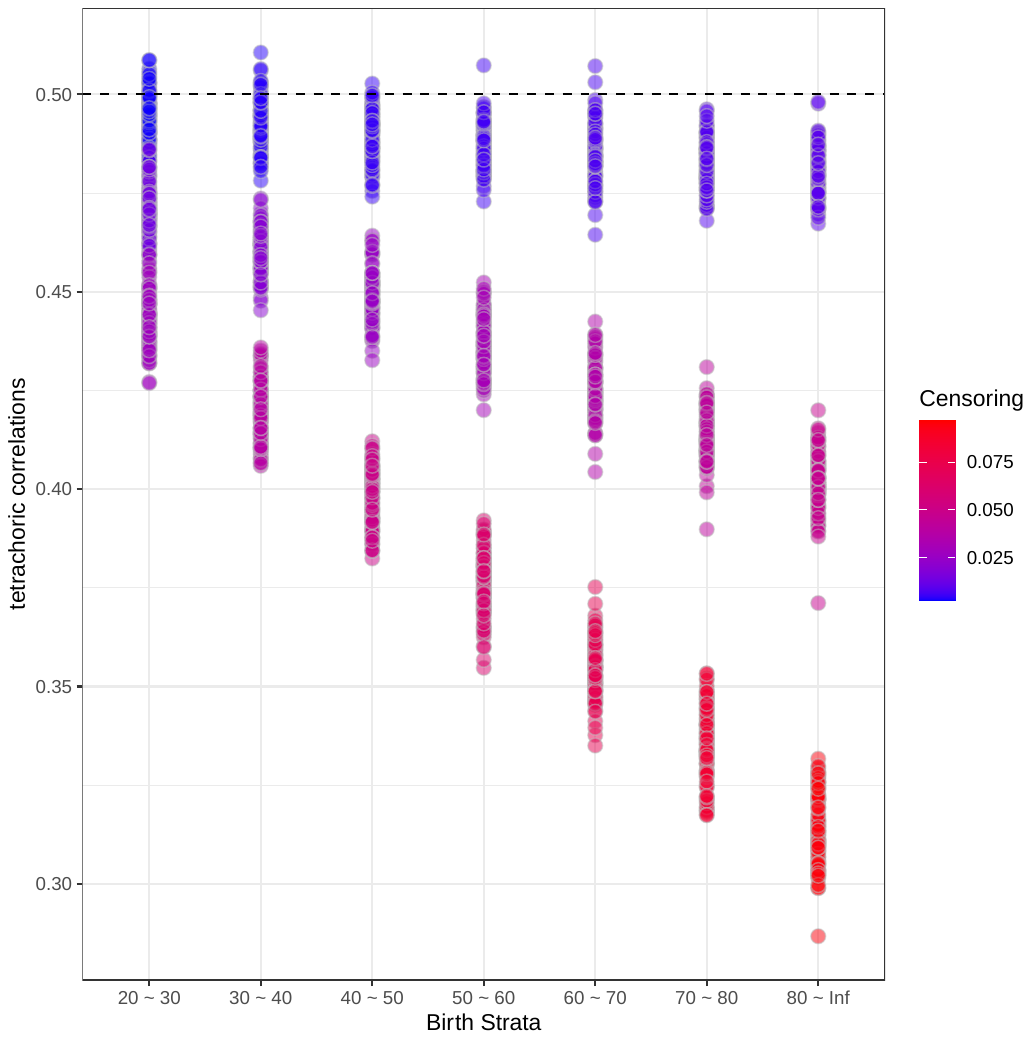

Supplementary Figure 3. Simulations on the impact of censoring on the estimations of AM with tetrachoric correlation. The total cohort size is fixed at 10,000 pairs while the age of the individuals are assigned as uniformly random. In this scenario, the prevalence is fixed at 0.2 and the true AM is fixed at 0.5. We varied three different levels of censoring as functions of age, resulting in differential censoring over birth strata. Each parameter scenario is repeated 100 times. The result shows that the point estimate of the tetrachoric correlations is biased downward when the censoring exists. The estimated AM becomes always lower than the true value, regardless of the strength of censoring.

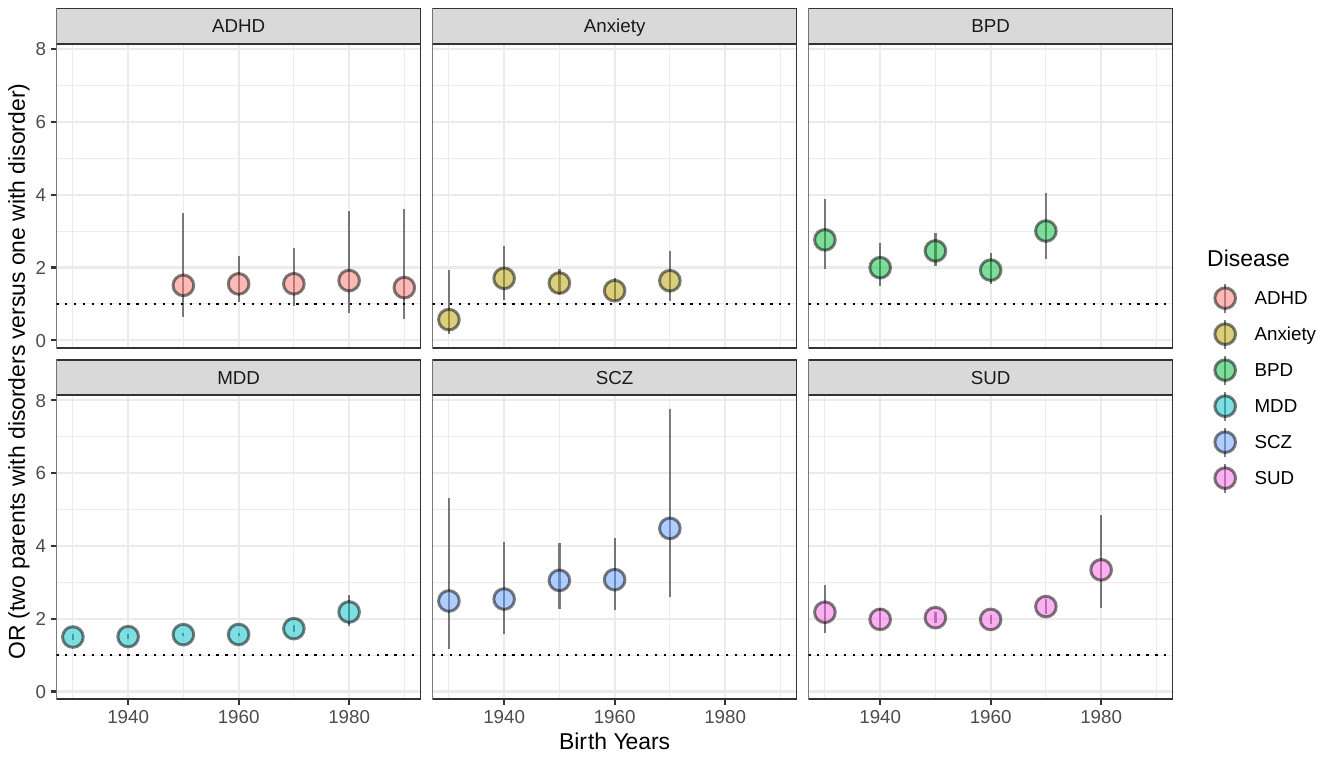

Supplementary Figure 4. Odds ratios of those who have two parents with disorders versus those who have only one parent with disorders, per parental birth strata. Error bars are 95% confidence intervals based on normal approximations of log(OR). ASD, AN, and OCD have limited number of parent-offspring pairs that preventing such analyses, hence not showing here.

Supplement Table 1. ICD code for nine psychiatric disorders.

| **Diseases** | **ICD 9** | **ICD 10** |
| --- | --- | --- |
| ADHD | 314 | F90 |
| SCZ | 295.x | F20, F25 |
| Anxiety | 300.0 | F40, F41 |
| ASD | 299.x | F84 |
| BPD | 296.0-296.1, 296.4-296.8 | F30, F31, F34.0 |
| MDD | 296.2x, 296.3x, 300.4, 311 | F32, F33, F34.1 |
| SUD | 291, 292, 303.0, 303.9, 304, 305 | F10-F19 |
| OCD | 300.3 | F42 |
| AN | 307.1 | F50.0 |

Supplement Table 2. Characteristics of Taiwanese cohort, Female probands and their matched controls.

TW_N_Summary_Female_Probands.csv

Supplement Table 3. Characteristics of Taiwanese cohort, Male probands and their matched controls.

TW_N_Summary_Male_Probands.csv

Supplement Table 4. Characteristics of Danish cohort

DK_Sample_Summary.csv

Supplement Table 5. Characteristics of Swedish cohort

SWD_Sample_Summary.csv

Supplement Table 6. GWAS summary statistics for nine psychiatric disorders

| **Trait** | **Data Source** | **Cases** | **Controls** | **Reference** |
| --- | --- | --- | --- | --- |
| **ADHD** | PGC | 19,099 | 34,194 | Demontis, D., et al. (2019). Discovery of the first genome-wide significant risk loci for attention deficit/hyperactivity disorder. Nature Genetics, 51(1), 63-75 |
| **SCZ** | PGC | 53,386 | 77,258 | The Schizophrenia Working Group of the Psychiatric Genomics Consortium, et al. (2020). Mapping genomic loci prioritises genes and implicates synaptic biology in schizophrenia. medRxiv 2020.09.12.20192922 |
| **Anxiety** | PGC | 5,540 | 11,770 | Otowa, T. et al. (2016). Meta-analysis of genome-wide association studies of anxiety disorders. Molecular Psychiatry 21, 1391-1397. |
| **ASD** | PGC | 18,381 | 27,969 | Grove, J. et al. (2019). Identification of common genetic risk variants for autism spectrum disorder. Nature Genetics, 51(3), 431-444 |
| **BPD** | PGC | 20,352 | 31,358 | Stahl, E. A., et al. (2019). Genome-wide association study identifies 30 loci associated with bipolar disorder. Nature Genetics, 51(5), 793-803 |
| **MDD** | PGC | 135,458 | 344,901 | Wray, N. R. et al. (2018). Genome-wide association analyses identify 44 risk variants and refine the genetic architecture of major depression. Nature Genetics, 50(5), 668-681. |
| **SUD** | PGC | 8,485 | 20,272 | Walters, R. K. et al. (2018). Transancestral GWAS of alcohol dependence reveals common genetic underpinnings with psychiatric disorders. Nature Neuroscience, 21(12), 1656-1699. |
| **OCD** | PGC | 2,688 | 7,037 | International Obsessive Compulsive Disorder Foundation Genetics Collaborative (IOCDF-GC) and OCD Collaborative Genetics Association Studies (OCGAS) et al., (2018). Revealing the complex genetic architecture of obsessivecompulsive disorder using meta-analysis. Molecular psychiatry, 23(5), 1181-1188. |
| **AN** | PGC | 16,992 | 55,525 | Watson, H. J., et al. (2019). Genome-wide association study identifies eight risk loci and implicates metabo-psychiatric origins for anorexia nervosa. Nature Genetics, 1207-1214. |
